# Risk of BA.5 infection in individuals exposed to prior SARS-CoV-2 variants

**DOI:** 10.1101/2022.07.27.22277602

**Authors:** João Malato, Ruy M. Ribeiro, Pedro Pinto Leite, Pedro Casaca, Eugénia Fernandes, Carlos Antunes, Válter R. Fonseca, Manuel Carmo Gomes, Luis Graca

## Abstract

The SARS-CoV-2 omicron BA.5 subvariant is progressively displacing earlier subvariants, BA.1 and BA.2, in many countries. One possible explanation is the ability of BA.5 to evade immune responses elicited by prior BA.1 and BA.2 infections. The impact of BA.1 infection on the risk of reinfection with BA.5 is a critical issue because adapted vaccines under current clinical development are based on BA.1.

We used the national Portuguese COVID-19 registry to analyze the risk of BA.5 infection in individuals without a documented infection or previously infected during periods of distinct variants’ predominance (Wuhan-Hu-1, alpha, delta, BA.1/BA.2). National predominance periods were established according to the national SARS-CoV-2 genetic surveillance data (when one variant represented >90% of the sample isolates).

We found that prior SARS-CoV-2 infection reduced the risk for BA.5 infection. The protection effectiveness, related to the uninfected group, for a first infection with Wuhan-Hu-1 was 52.9% (95% CI, 51.9 – 53.9%), for Alpha 54.9% (51.2 – 58.3%), for Delta 62.3% (61.4 – 63.3%), and for BA.1/BA.2 80.0% (79.7 – 80.2%).

The results ought to be interpreted in the context of breakthrough infections within a population with a very high vaccine coverage (>98% of the study population completed the primary vaccination series).

In conclusion, infection with BA.1/BA.2 reduces the risk for breakthrough infections with BA.5 in a highly vaccinated population. This finding is critical to appraise the current epidemiological situation and the development of adapted vaccines.

## Results and Discussion

In recent months, omicron (B.1.1.529) became the dominant SARS-CoV-2 variant, displaying some degree of immune evasion^1,2^. The initial omicron subvariants, BA.1 and BA.2, are being progressively displaced by BA.5 in many countries, possibly due to greater transmissibility and partial evasion of BA.1/BA.2-induced immunity^3,4^. The protection afforded by BA.1 towards BA.5 infection is critical as adapted vaccines under clinical trials are based on BA.1.

Portugal was one of the first countries affected by a BA.5 predominance. We used the national COVID-19 registry (*SINAVE*) to calculate the risk of BA.5 infection in individuals with documented infection with past variants, namely BA.1/BA.2. The registry includes all reported cases in the country, regardless of clinical presentation.

The national SARS-CoV-2 genetic surveillance identified periods when different variants represented >90% of the isolates^5^. We identified all individuals that had the first infection in periods of dominance of each variant, and a second infection in the period of BA.5 dominance (Figure 1A). We pooled BA.1 and BA.2 because of the slow transition between the two subvariants in the population. Finally, we calculated the risk of BA.5 infection for the population that did not have any documented infection before the BA.5 dominance (June 1st).

**Figure 1.**
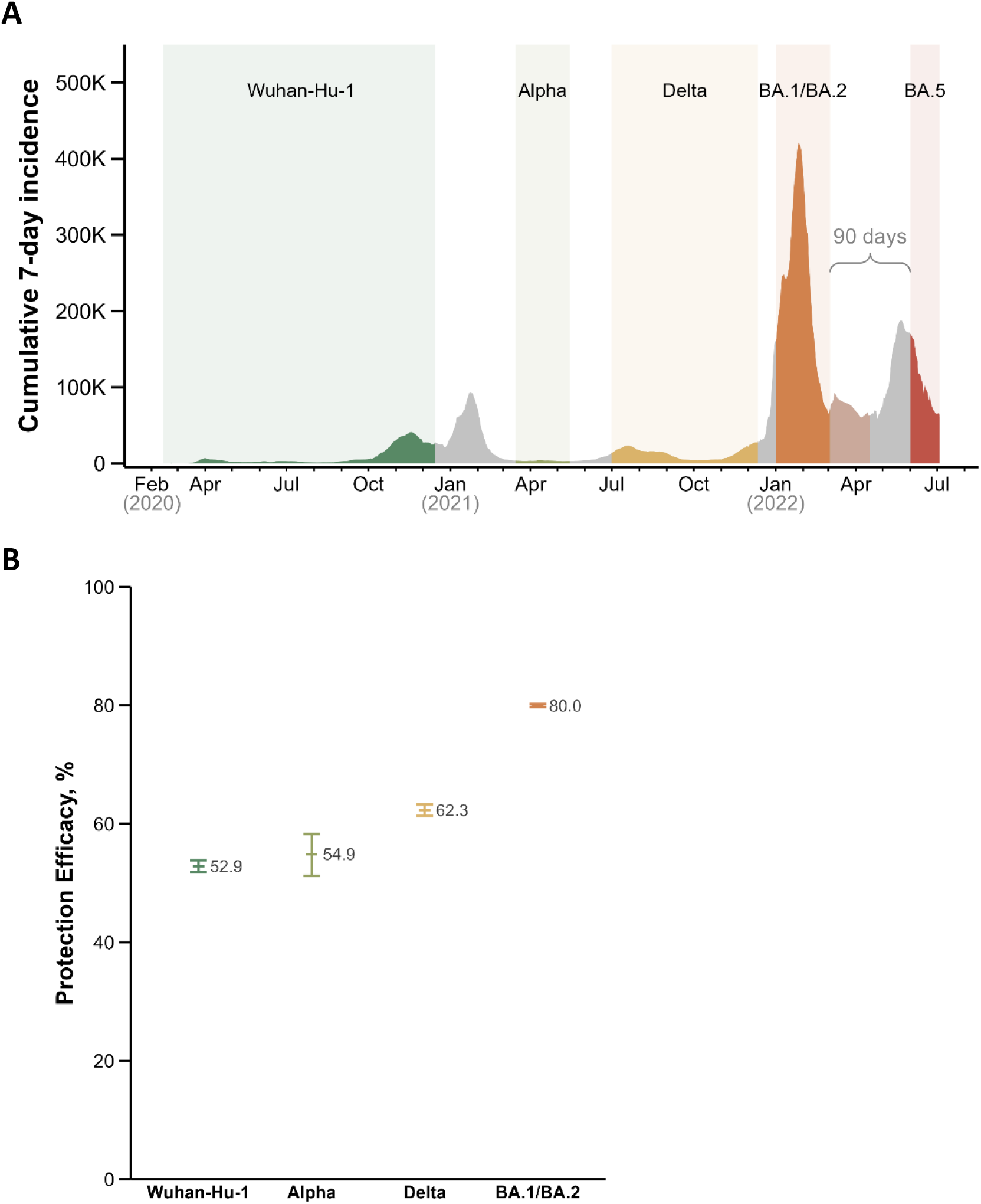
Protective effect of previous SARS-CoV-2 infection on infection with omicron BA.5 subvariant. **A**. We identified the periods (in different colors) where one variant was represented in >90% of sample isolates (data from the national SARS-CoV-2 genetic diversity surveillance^5^). The periods in grey represent times when more than one variant was in circulation. Given the relatively slow transition between BA.1 and BA.2 dominance we pooled BA.1 and BA.2 in the analysis. We did not include anyone infected in the 90 days before BA.5 dominance. **B**. Protection efficacy against infection during the period of BA.5 dominance (from June 1st) for individuals with one infection in the periods of dominance of different variants, as represented in (A), in relation to individuals without any documented infection until June 1st. Individuals with two infections before June 1st were not included in the study.

We found that prior SARS-CoV-2 infection has a protective effect regarding BA.5 infection (Table 1, Figure 1B), and this protection is maximal for prior infection with BA.1/BA.2.

**Table 1.** Risk of omicron BA.5 infection according to previous infection history. We included in the study all the population 12 years and older. Under “1st infection” is the number of individuals at risk for a second infection by BA.5 (i.e., all individuals with a second infection before June 1st were excluded). Reinfections were defined as two positive tests by the same individual more than 90 days apart. Note that the risk is dependent on the epidemic situation in Portugal from June 1st to the end of the study (July 4th), affecting all groups equally. OR, odds ratio; CI, confidence interval.

|  | Uninfected on<br>June 1<sup>st</sup> | 1<sup>st</sup><br>infection | BA.5<br>infection | Absolute<br>Risk | OR (95% CI) | Protection Efficacy,<br>% (95% CI) |
| --- | --- | --- | --- | --- | --- | --- |
| Uninfected | 5 328 287 | – | 367 783 | 0.069 | – | – |
| Wuhan-Hu-1 | – | 267 448 | 9 031 | 0.034 | 0.471 (0.461, 0.481) | 52.9 (51.9, 53.9) |
| Alpha | – | 20 004 | 647 | 0.032 | 0.451 (0.417, 0.488) | 54.9 (51.2, 58.3) |
| Delta | – | 232 831 | 6 329 | 0.027 | 0.377 (0.367, 0.386) | 62.3 (61.4, 63.3) |
| BA.1/BA.2 | – | 1 557 635 | 22 793 | 0.015 | 0.200 (0.198, 0.203) | 80.0 (79.7, 80.2) |

These data should be considered in the context of breakthrough infections in a highly vaccinated population, given that in Portugal over 98% of the study population completed the primary vaccination series before 2022.

One limitation is the putative effect of immune waning in a population with hybrid immunity (infection + vaccine). We found that BA.1/BA.2 infection in vaccinated individuals provides higher protection against BA.5, in line with a recent report with a test-negative design^6^. However, BA.1/BA.2 infections occurred closer to the period of BA.5 dominance. There is a perception that protection afforded by prior BA.1/BA.2 infection is very low, given the high number of BA.5 infections in individuals with prior BA.1/BA.2 infection. Our data indicates that this perception likely is a consequence of the larger pool of individuals with BA.1/BA.2 infection, and it is not supported by the data.

Overall, we found that breakthrough infections with the BA.5 subvariant are less likely in individuals with a prior SARS-CoV-2 infection history in a highly vaccinated population, especially for prior BA.1/BA.2 infection.

In this study, we divided the Portuguese epidemic curve into time strata, each characterized by the dominance of one of the SARS-CoV-2 variants or subvariants. We then estimated the risk of infection during the Omicron/BA.5 period, for naüve individuals and for those infected in every stratum at least 90 days before the BA.5 period.

We found that, among a highly vaccinated population, the risk of infection by BA.5 was greater for individuals who had no documented infection, as compared to those who acquired hybrid immunity following a SARS-CoV-2 infection. We also found those infected with the Omicron BA.1/BA.2 subvariants were less at risk of a BA.5 infection than individuals infected with a previous variant. This heightened protection following BA.1/BA.2 infection can be due to the induction of a more effective immune protection towards BA.5 and/or to the shorter time elapsed between infection and exposure to the Omicron BA.5 subvariant.

We used a registry-based study that lacks the precision of a test-negative design^7^. However, the large number of cases that we used, resorting to the entire resident population of Portugal over 12 years old, led to a risk estimate for individuals with prior BA.1/BA.2 infection extremely close to an estimate from Qatar based on a test-negative design^6^. The effectiveness of protection of BA.1/BA.2 infection against BA.4/BA.5 was calculated in the study from Qatar as 79.7% (95% CI: 74.3 – 83.9%), in striking agreement with our data: 80.0% (79.7 – 80.2%). Our estimate for effectiveness of protection for individuals infected with SARS-CoV-2 pre-omicron variants was higher than in the study from Qatar, but not too dissimilar from a recent systematic review and meta-analysis that evaluated protection against omicron^8^.

The risk of infection is influenced by the immune status following vaccination and infection and by the time elapsed between the first infection and the BA.5 period due to immune waning. Both issues may be interlocked: in situations where some degree of immune evasion is present, the waning of protection appears to be more rapid. Omicron subvariants are known to differ from pre-omicron variants in having a higher capacity to evade humoral immunity^1–4^. It was demonstrated for immune responses directed towards SARS-CoV-2 that the decline of neutralizing antibodies over time appears to follow a similar slope^9^. Hence, partial immune evasion can lead to a lower titre of neutralizing antibodies that, with a similar slope of decline, will tend to be faster in reaching a level of neutralizing antibodies unable to sustain protection from infection. The relationship between immune evasion and more rapid waning was well illustrated for omicron subvariants^10^. Still, the protection that we are calculating is the protection relevant for public health since infection with previous variants occurred at different times in the past. In any case, for public health, the critical information is related to the protection against BA.5 observed with individuals infected with previous variants at the time they were in circulation, as it represents the current situation.

In addition, for the interval that we studied (3 – 5 months after BA.1/BA.2 infection), it is important that the protection conferred by this hybrid immunity remains very significant compared with people vaccinated and without documented infection, or with infection with pre-omicron variants. Indeed, our results are consistent with other studies showing that omicron subvariants are superior to pre-omicron variants in leading to greater protection against other omicron subvariants^6–8,11^. Subsequent studies, with longer follow-up, will be necessary to establish the extent of a putative (but likely) effect of immune waning in this hybrid immunity.

A possible confounder to our results is the presence of undocumented infections among the “uninfected” group of individuals (without a positive test in the registry). A national serologic survey was performed by the National Health Institute Ricardo Jorge before vaccination and at different times following vaccination (using SARS-CoV-2 specific anti-N IgG)^12^. With data from the last national serologic survey, we estimated the population with serologic evidence of SARS-CoV-2 infection that was not detected by testing (i.e., that was not present in the COVID-19 registry) as 29.2% of the notified infections^12^.

We modeled the impact of the presence within the “uninfected” group of unreported infected individuals (in a frequency coherent with the national serologic survey). For this, we calculated the number of all SARS-CoV-2 infections before June 1^st^ and multiplied this number by 0.292 to obtain the number of undetected cases (605,944; Table 2). This number of undetected cases was subtracted from the “uninfected”. Next, we removed the reinfections with BA.5 assigned to “uninfected” for the 605,944 individuals that were, in fact, infected. For this, we used the absolute risk calculated for the entire population of individuals that had an infection until 90 days before June 1^st^ 2022 – this strategy considers the relative contribution of the number of infections with the distinct variants (and even periods when no variant was dominant) and their risk. The absolute risk was calculated as 0.020. Hence, the number of BA.5 infections that was excluded from the previously uninfected was 12,208. It is not necessary to distribute the new cases of infection for the different groups of variants as these extra cases of infection, and their respective reinfections would not change the relative risks (since this distribution would be proportional across all groups).

**Table 2.** Risk of omicron BA.5 infection according to previous infection history, considering an estimate of unreported cases of infection. We used data from the national COVID-19 serologic survey that estimated that an additional 29.2% of cases of infection which were not reported^5^. We calculated the overall absolute risk of reinfection during the period of BA.5 dominance for all the infections as 0.020. The values under “Estimated true uninfected” were obtained by subtracting the number of unreported infections and BA.5 reinfections (assuming a 0.020 absolute risk.). OR, odds ratio; CI, confidence interval.

|  | Uninfected<br>June 1<sup>st</sup> 22 | 1<sup>st</sup> infection | BA.5<br>infection | Absolute<br>Risk | OR (95% CI) | Protection Efficacy, %<br>(95% CI) |
| --- | --- | --- | --- | --- | --- | --- |
| Infection<br>non-reported | – | (605 944) | (12 208) | 0.020 | – | – |
| Estimated<br>true uninfected | 4 722 343 | – | 355 575 | 0.075 | – | – |
| Wuhan-Hu-1 | – | 267 448 | 9 031 | 0.034 | 0.429<br>(0.420, 0.438) | 57.1<br>(56.2, 58.0) |
| Alpha | – | 20 004 | 647 | 0.032 | 0.410<br>(0.380, 0.444) | 59.0<br>(55.6, 62.0) |
| Delta | – | 232 831 | 6 329 | 0.027 | 0.343<br>(0.335, 0.352) | 65.7<br>(64.8, 66.5) |
| BA.1/BA.2 | – | 1 557 635 | 22 793 | 0.015 | 0.182<br>(0.180, 0.185) | 81.8<br>(81.5, 82.0) |

Table 2 shows that including the unreported positive cases led to a slight increase in the effectiveness of protection (for BA.1/BA.2 from 80% to 82%) (Figure 2A). We further explored this sensitivity by considering that unreported cases comprised 20% or 40% of infected cases and redoing the protection calculations (Figure 2B). In all situations, consideration of unreported cases lead to an increase in the risk of primary BA.5 infection in the previously uninfected group, and thus an increase in the relative protection of infection with a prior SARS-CoV-2 variant.

**Figure 2.**
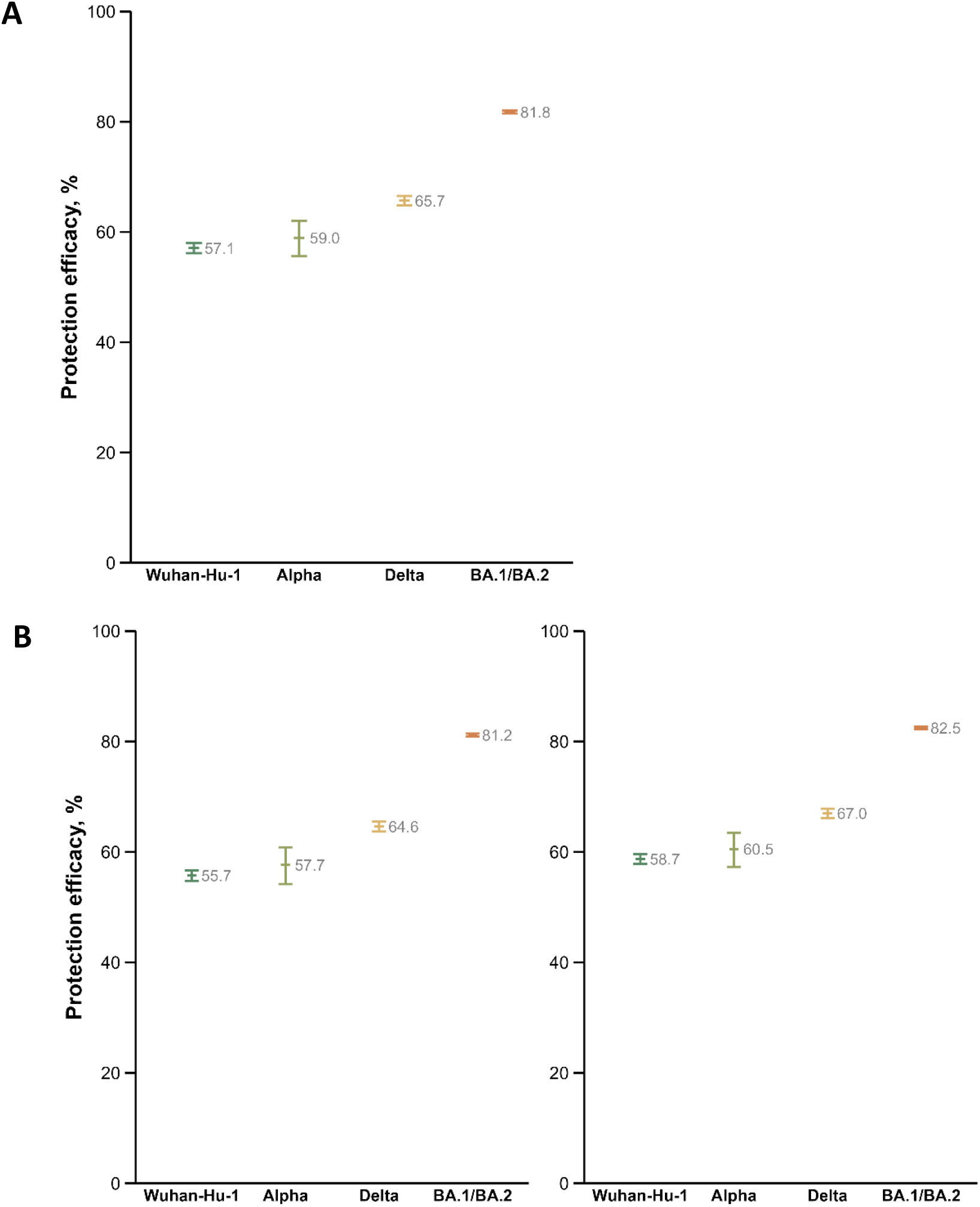
Estimates of the impact of unreported cases of infection among the population absent from the national COVID-19 registry. The most recent national COVID-19 serologic survey estimated that an additional 29.2% of cases of infection were not notified^5^. This figure was calculated based on the seroprevalence of SARS-CoV-2 anti-N IgG in the population. (A) We calculated the protection efficacy for prior infection with different SARS-CoV-2 variants in relation to the uninfected population, after correction with the unreported cases of infection (removed from the “uninfected” group). (B) To further explore the method’s sensitivity, we calculated protection efficacy for scenarios where the unreported cases were 20% (left) and 40% (right) of all notified infections.

We note that we could also consider unreported cases in each of the groups with previous infection with different variants. This would increase by some percentage the number of first infections with Wuhan-Hu-1, Alpha, Delta, and BA.1/BA.2, leading to a lower absolute risk of secondary infection with BA.5 in these groups. Relative to no previous infection, this would increase the protection of having had an infection before. Finally, the number of cases of BA.5 could also be larger, because of unreported cases. However, any increase in these numbers would affect all groups (uninfected and previously infected) in a similar proportion, because we are considering BA.5 infections over the same time frame in all cases. Thus, putative unreported cases in BA.5 would have minimal impact on the relative protection of the previous infections. We assumed that the frequency of unreported cases of infection remained relatively constant over the periods of dominance with different variants. It is very likely that the frequency of unreported cases increased during the period of BA.5 dominance, given the change in the testing policy in Portugal. However, for the reasons given above, the change in this period is expected to affect all groups similarly. It is anticipated that more subtle changes in the frequency of unreported cases occurred over the previous periods, given the maintenance of public health policies, namely regarding testing. This is also confirmed by the relatively constant frequency of estimated unreported cases in the three periods evaluated by the national COVID-19 serologic survey^12^.

The sensitivity analysis with a variable percentage of false negatives confirmed that the main conclusions are similar despite this issue. The experimental design does not account, however, for characteristics that may differ between infected and uninfected: some high-risk occupations, lifestyles, and living environments may place individuals consistently at higher risk, that were likely to be infected earlier in the pandemic. Nevertheless, our data is coherent with several other reports consistently showing greater protection of omicron subvariants towards reinfection with other omicron subvariants^6,11^.

The popular perception that BA.1/BA.2 infection cannot protect against BA.5 reinfection, a consequence of the observation that many people infected in the first two months of 2022 (BA.1/BA.2 dominance) were reinfected by BA.5 is a perception error. Probably explained by the very high number of infections caused by BA.1/BA.2. In Portugal, there were 1.97 million documented infections in January-February/2022 alone, which compares with 1.42 million during the entire epidemic before.

Our results, together with a similar report^6^, show that infection with BA.1/BA.2 of a population mostly vaccinated provided significant protection against BA.5 reinfection. At a time when BA.1-based adapted vaccines are under clinical development, this information is of great importance.

## Methods

### Participant selection

The population included in the study was all Portuguese residents aged 12 years and older, obtained from the National Census 2021 ^13^.

We used the national COVID-19 registry (*SINAVE*) to obtain information on all notified cases of infection, irrespective of clinical presentation. The “uninfected” population was defined as the population over 12 years of age without a documented infection in the registry. The number of uninfected people in June 1^st^ 2022 (the start of the study period) was 5 328 287, representing 57% of the Portuguese population over 12.

The data available in the national COVID-19 registry (*SINAVE*) only include cases of tests (PCR tests and rapid antigen tests) performed by healthcare workers in accredited diagnostic facilities. Testing by an accredited facility is a requisite for access to social security compensation for days of isolation – this is a reason for the comprehensiveness of the registry and the exclusive inclusion of validated tests. Only tests performing above the EU-defined minimum for test sensitivity and specificity are used in Portugal. Furthermore, until recently, Portugal had a wide and mandatory testing policy, requiring the presentation of tests for access to several locations, even for vaccinated people (namely, access to entertainment, sports, or healthcare venues).

It is anticipated that the population we classified as “uninfected” contained individuals with a prior unnoticed infection. This issue is discussed below.

We used the national SARS-CoV-2 genetic surveillance database^5^ to identify periods when different variants represented >90% of the sample isolates, as also used in other studies^6^. With this information, we identified the individuals who were infected in the period of dominance of each variant (Wuhan-Hu-1, Alpha, Delta, BA.1/BA.2, BA.5; Table 3). We pooled the BA.1 and BA.2 infections, given the slow transition between the period of dominance of these two subvariants.

**Table 3.** Periods of dominance of the different SARS-CoV-2 variants and omicron subvariants in Portugal. In each of the periods, the variant or subvariant was represented in >90% of sample isolates of the Portuguese residents (data from the national SARS-CoV-2 genetic diversity surveillance^2^). * We used a dataset until July 4^th^ 2022.

| Variant or<br>Omicron suvariant | Start | End |
| --- | --- | --- |
| Wuhan-Hu-1 | 14 Feb 20 | 15 Dec 20 |
| Alpha | 15 Mar 21 | 15 May 21 |
| Delta | 1 jul 21 | 12 Dec 21 |
| BA.1 | 1 Jan 22 | 6 Feb 22 |
| BA.2 | 27 Mar 22 | 17 Apr 22 |
| BA.5 | 1 Jun 22 | present* |

We excluded from the analyses all individuals who had more than one infection before June 1^st^ (see the flowchart in Figure 3).

**Figure 3.**
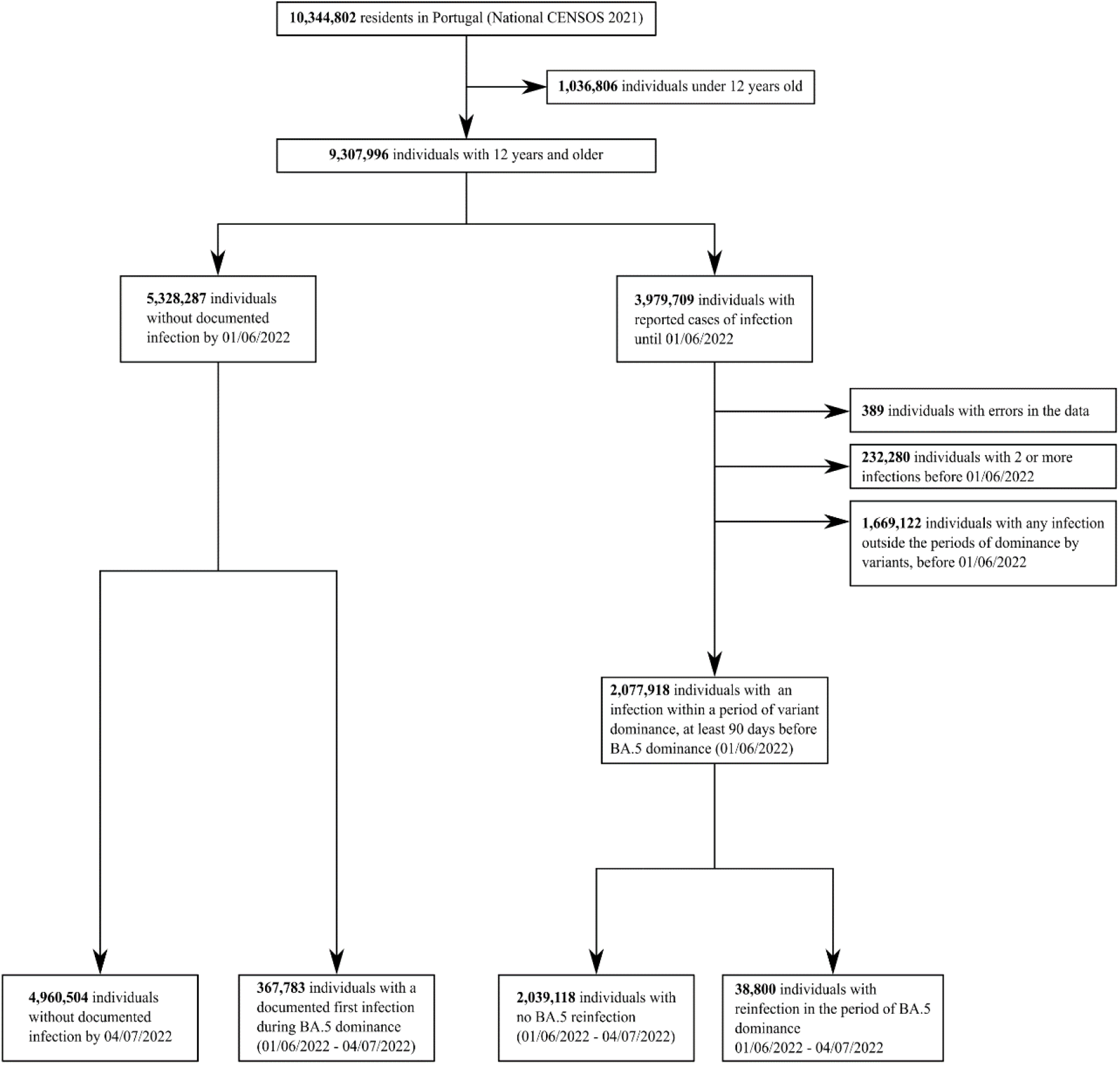
Flowchart describing the population selection.

Reinfection was defined as two positive tests in the same individual, at least 90 days apart^14^. Consequently, all cases of infection in the 90 days before the start of BA.5 dominance were not included, as these would not classify as “in risk of reinfection” for the entire duration of the test period under the definition above.

In summary, the population included in the study comprises: (1) All individuals resident in Portugal aged 12 years and older without a documented infection until June 1^st^ 2022; (2) All individuals resident in Portugal aged 12 years and older with a single documented infection before June 1^st^, when this infection occurred during periods of clear dominance (>90% of cases) of the different variants, but not in the 90 days before June 1^st^.

### Vaccination coverage

The vaccine coverage with the primary vaccination series in the Portuguese residents over 12 years was >98% by the end of 2021. The primary series of the vaccination campaign used EU/EMA-authorized vaccines: Comirnaty (Pfizer/BioNTech), 69%; Spikevax (Moderna), 12%; Vaxzevria (AstraZeneca), 13%; and Janssen 6%.

At the start of the BA.5 period of dominance (June 1^st^), the coverage with the first booster was 82%, exclusively using mRNA vaccines (77% Comirnaty and 23% Vaxzevria). A second booster was not yet in use except for a highly specific (and small) population of patients with severe immunosuppression.

### Statistics

We calculated the absolute risk of BA.5 infection for people with different prior history of infection, as the number of cases of BA.5 in the period under consideration over the number of people at risk in each group (uninfected, and single previous infection with Wuhan-Hu-1, Alpha, Delta or BA.1/BA.2). We also used these numbers to calculate odds ratio (OR), that is the ratio of the odds of the previous infection (for different variants) to previously uninfected in the BA.5 infected people to these odds in people not infected with BA.5. Protection efficacy was estimated, in percentage, as (1-OR) x 100%. Confidence intervals for the OR were calculated using the normal approximation method.

Further, we performed a sensitivity analysis for the possibility of undiagnosed cases of SARS-CoV-2 in the uninfected population. We used data from the National Serological Panel (from November 2021, assaying the presence of antibodies against the N protein to exclude vaccination seropositivity) to infer that, at that time, there were 29.2% more people who had been infected with SARS-CoV-2 than officially reported^12^. We calculated how many more infections this would correspond to on June 1^st^, 2022, and removed these infected unreported cases from our uninfected population. We estimated that 2% of these would have also been reinfected with BA.5 during the period of interest (being 0.020 the absolute risk for reinfection for the global population with a prior infection). We thus removed that number from the BA.5 infected cases (see Table 2). With these new values, we recalculated the absolute risk of BA.5 in the previously uninfected and the OR. To further ascertain the sensitivity of our results, we also tested the assumtions that maybe there were only 20% or up to 40% more people who had been infected than officially reported and repeated the calculations described above.

## Data Availability

All data produced in the present study are available upon reasonable request to the authors.

https://insaflu.insa.pt/covid19

